# Airborne Pathogens in a Heterogeneous World: Superspreading & Mitigation

**DOI:** 10.1101/2020.10.24.20218784

**Authors:** Julius B. Kirkegaard, Joachim Mathiesen, Kim Sneppen

## Abstract

Epidemics are regularly associated with reports of superspreading: single individuals infecting many others. How do we determine if such events are due to people inherently being biological superspreaders or simply due to random chance? We present an analytically solvable model for airborne diseases which reveal the spreading statistics of epidemics in socio-spatial heterogeneous spaces and provide a baseline to which data may be compared. In contrast to classical SIR models, we explicitly model social events where airborne pathogen transmission allows a single individual to infect many simultaneously, a key feature that generates distinctive output statistics. We find that diseases that have a short duration of high infectiousness can give extreme statistics such as 20 % infecting more than 80 %, depending on the socio-spatial heterogeneity. Quantifying this by a distribution over sizes of social gatherings, tracking data of social proximity for university students suggest that this can be a approximated by a power law. Finally, we study mitigation efforts applied to our model. We find that the effect of banning large gatherings works equally well for diseases with any duration of infectiousness, but depends strongly on socio-spatial heterogeneity.

## INTRODUCTION

The statistics of an on-going epidemic depend on a number of factors. Most directly: How easily is it transmitted? And how long are individuals affected and infectious? Scientific papers and news paper articles alike tend to summarize the intensity of epidemics in a single number, *R*_0_. This *basic reproduction number* is a measure of the *average* number of individuals an infected patient will successfully transmit the disease to. It has become evident, however, that in many epidemics most people who are infected do not themselves infect even a single other individual. The trajectories of the such epidemics are instead driven by fewer people who infect many [1]. As a statistical phenomenon, this can be captured by modelling individuals with a varying reproduction number [2].

But what is the cause of this dispersion? The simplest explanation is perhaps that of biological variation. Some people might inherently be *superspreaders*, and for the recent spreading of Covid-19 [3–5] there is mounting evidence for this [6]. However, other sources of heterogenity could also explain much of the observed super-spreading [7], including heterogeneous social interactions [8, 9], which has previously been studied by simulating epidemic spreading on heterogeneous networks [10–12]. Typical models use variations of the SIR (Susceptible-Infected-Recovered) model [13] which, even in the case of modelling spread on heterogeneous network, has the property that during each ‘time step’ each individual only interacts with one other. This implicitly assumes a method of spreading that involves touch.

Here, we consider a model for disease propagation in which an infectious individual can infect several people at the same time, which will be the case when the method of transmission is through air. This change has consequences not only for the average infection rate, but also for the variation in the number of secondary infections. We consider people with equal infectiousness and expect superspreading events to occur when infectivity co-occur with a large gathering of people. We explicitly model social activity by assuming that people participate in social events of different sizes assigning a risk of infection that increases at crowded places. We derive the spreading statistics caused by this heterogeneity alone, and predict superspreading when the infectious period is short.

## MODEL

In our model world people move from location to location, and infected people are imagined to spread a cloud of virus that can infect everybody within some distance set by the property of the disease, as illustrated in Fig. 1. We consider time discrete, and assume that at each timestep people move to a new location. The duration of infectiousness is parametrized by a number *M*, which counts the number of locations a person visits while being infectious. Thus a very short period with high infectivity is modeled by *M* = 1, while large *M* corresponds to a disease with a prolonged infectious state.

**FIG. 1.**
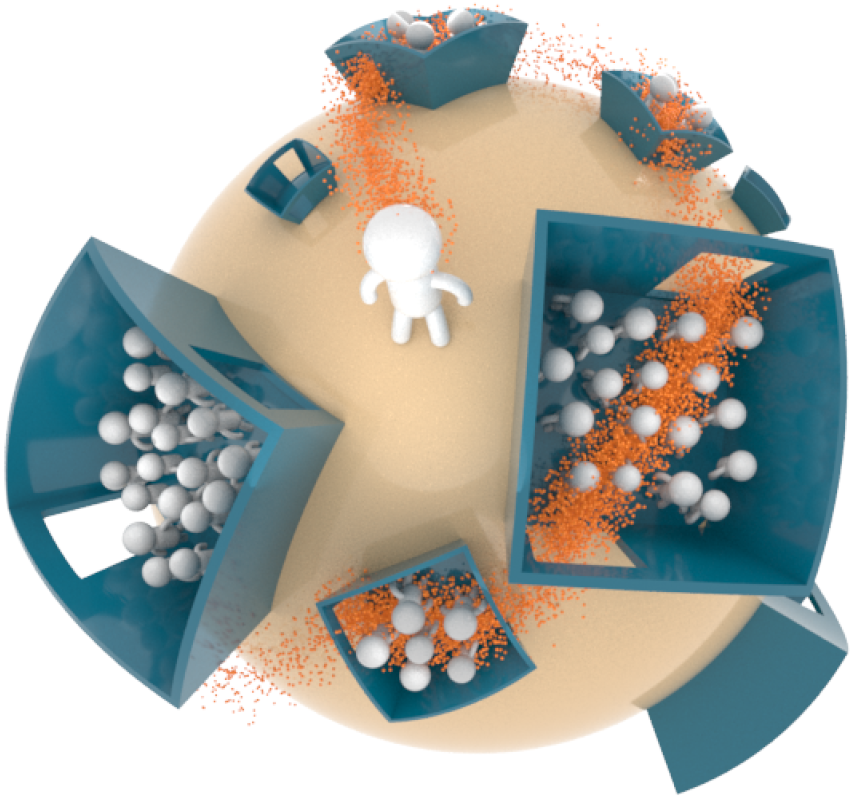
Illustration of model world. An infectious person visits *M* places while being infectious, at each visit spreading airborne pathogens that can infect many. The number of people that can potentially be infected at each location depends both on the size of the location and the range of spread of the airborne pathogens.

Our social world model consists of visits to locations, and is quantified by the size distribution over these social gatherings. Consider first the case in which an infected individual at each time step visits a location where the number of people is exponentially distributed. For fixed *R*_0_, the total the number of secondary infections will then follow a negative binomial distribution with parameter (1 + *R*_0_*/M*)^−1^, independent of the shape of the exponential distribution (see SI). Thus the statistics will at most be marginally overdispersed, and in particular we find that the 20 % most infectious individuals will at most infect 65 % for *R*_0_ ≥ 1.0.

However, many social events are much larger than the family size events that dominate our daily life. Thus we assume a fat-tailed probability density function for the chance of being at a location in which one on average could infect up to *n* other people,

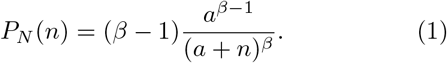

Here the exponent *β >* 2 determines the broadness of the distribution: For small *β* people will often visit locations where they interact with large crowds. The distribution corresponds to picking a random person and asking how many other people the airborne particles from this person will reach on average. This will typically be lower than the total number of people at the location. Further, this distribution is marginalized over time in the sense that it should be thought to include rare events that people go to perhaps just once a month, etc.

The number of people an individual infects at a location *Z*_*i*_ we take to follow Poisson statistics, *Z*_*i*_|*N*_*i*_ ∼ Pois(*αN*_*i*_), where *α* is the overall infectiousness of the disease and *N*_*i*_ is a random number drawn from *P*_*N*_. This effectively means that a single person can infect many in a single timestep of the model, which is in line with how airborne pathogens are believed to spread [14].

We derive the solution to our model in the methods section. We note that while we discretize time, a qualitatively similar model can be developed with time continuous and for which the duration of events vary.

## RESULTS

### Superspreading

We consider epidemics of known basic reproduction number *R*_0_. In our model this average infection number reads

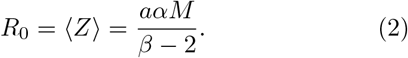

To compare varying values of *β* and *M*, we choose *α* such that *R*_0_ is fixed. Naturally, increasing the number of events (*M*), i.e. the duration of the infectious stage, the infectivity per event has to decrease in order to keep *R*_0_ fixed. Note how, as *β* → 2, the infectiousness must be scaled to zero, *α* → 0 reflecting the fact that infections become dominated by extremely large events. Thus for *β* → 2 the epidemic is driven solely by extreme super-spreading events.

The value of *M*, which is defined by the disease, and may differ widely between diseases with equal *R*_0_, has a crucial effect on the offspring distribution, as shown in Fig. 2. As *M* becomes small, to keep *R*_0_ fixed, *α* must be increased, and thus the chance of infecting a lot in a single location goes up. So a disease with a short time of infectiousness will be more prone to yield superspreading events, while long infectiousness gives statistics similar to a homogeneously spreading disease.

**FIG. 2.**
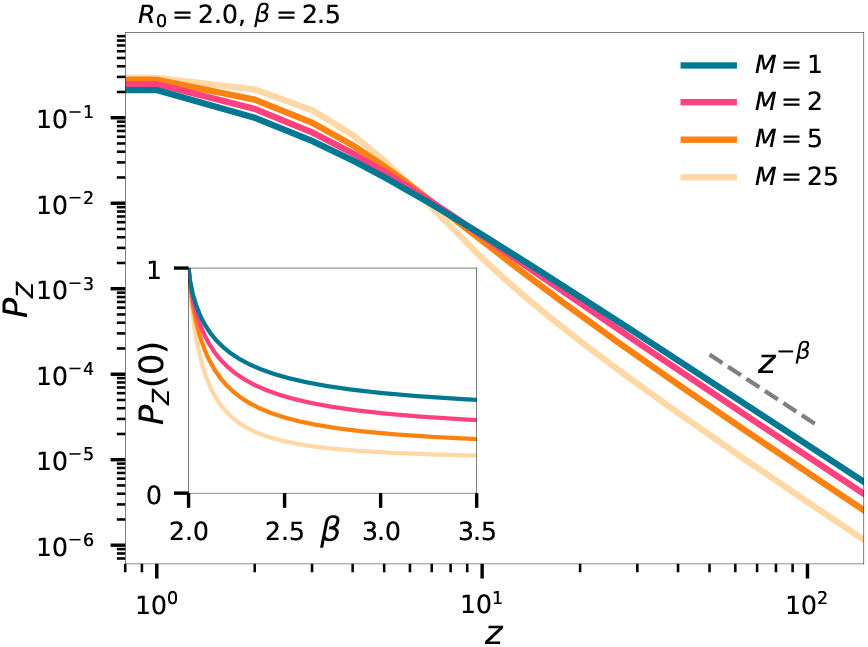
Offspring statistics. The distribution of *Z*, the number of individuals an infected individual will infect. Inset shows the probability that an individual will infect precisely zero. In all plots we take *a* = 3.

As a concrete example, we find from Fig. 2 that the probability of a person to infect 100 others is a full order of magnitude smaller with *M* = 25 compared to *M* = 1. The inset of Fig. 2 shows that as *M* is decreased, the chance of infecting exactly zero increases. The simple logic is that when a few is infecting many, there must be many who infect none. We note that in practice the effective *M* will typically be smaller than the duration of infectiousness if people are aware of the disease and its symptoms and decide to self-isolate. In this case, *M* should reflect the amount of time spent being infectious while on a normal daily routine and not the inherent duration of infectiousness of the disease. Thus an airborne diseases with short pre-symptomatic infectivity may likely be driven by superspreading events.

We present in Fig. 3 mobility tracking data of ∼650 students at a Danish university [details in methods section]. This data allow us to determine the average time that people spend at a location. We estimate this by calculating the distribution of time *τ* spent consecutively together for all pairs of students. The inset of Fig. 3(a) shows that this approximately follows an exponential distribution with characteristic time *τ*_0_ ∼ 1.6 hours. The fit does not match the data for small *τ*, where students simply passing one another, and not actually staying nearby one another, bias the data. If we denote the duration of infectiousness of the disease by *T*, and assume that the disease only spreads for meetings longer than a certain duration, one can approximate *M* ∼ *T/τ*_0_. Taking into account that for the student data presented here, time spent alone (such as sleeping) is not captured, we get an estimate of *M* in the range of 5 – 10 for *T* ∼ 24 hours.

**FIG. 3.**
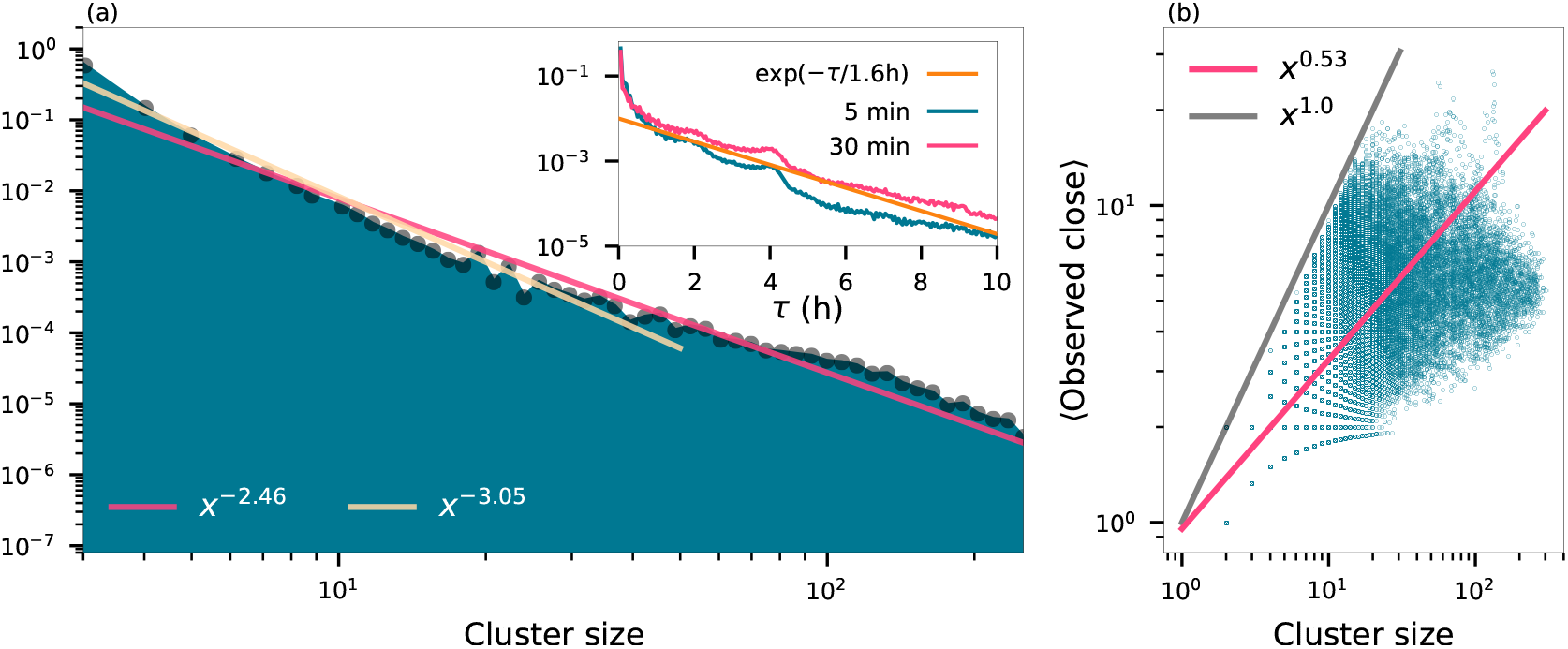
Statistics of social interaction networks generated from proximity data. Proximity is determined using Bluetooth by an app installed on smartphones distributed to a group of students at a Danish university (*N* ∼ 650). (a) The proximity data are split in one hour windows and in each window we identify the connected components of the social interaction network and plot the distribution of cluster sizes. Power law fits over the entire range of clusters gives ∼ *x*^−2.5^, whereas restricting the range to smaller cluster gives ∼ *x*^−3^. Inset shows the distribution of the duration of time that people spent together. Blue curve allows gaps in the data of 5 minutes and pink allows 30 minute gaps [see methods]. Fitting these curves with ∼exp(− *τ/τ*_0_) for times *>* 30 minutes yields characteristic times *τ*_0_ ≈ 1.5 hours for the blue curve and *τ*_0_ ≈ 1.7 for the pink curve. (b) For each cluster, the plot shows the average number of students that had a strong Bluetooth signal with one another indicating proximity; ≲ meters. A power law fit of the data yields ∼ *x*^0.5^.

The other crucial parameter in our model is the exponent *β*. It is well-established that the sizes of, for example, cities or companies approximately follow a Zipf distribution ∼*x*^−2^ asymptotically [15, 16]. Although firm sizes are indicative of places people visit, we expect such a distribution to be steeper in general. Further, the distribution will vary from country to country, city to city, and neighbourhood to neighbourhood. As a concrete example we employ the mobility tracking data, for which we find that the sizes of groups [Fig. 3(a)] that students gather in follow a power law distribution with exponent in the range of ∼ 2.5 – 3.0.

The number of potential infections at a location depends on the range and suspension time of the airborne pathogens. Although the tracking data is limited in size, and thus exponential cutoffs are quite small, we can nonetheless calculate the average number of people that are within short range of one another within each cluster. Fig. 3(b) shows that in a cluster of *x* people, each person in on average close to 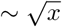 other people.

In the general case of a location size distribution, which asymptotically goes like ∼*x*^−*γ*^, the chance of picking a person at a location of size *n* must then scale as ∼*x*^−(*γ*−1)^. *P*_*N*_ describes the average number of people one interacts with at a location. If we assume that these *x* people are distributed in a two-dimensional space, and that during the visit a person walks in a few straight lines through this space, there will of the order of 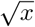 interactions, precisely as observed in the tracking data. Transforming we find *P*_*n*_ ∼ *n*^−2(*γ*−1)+1^. Thus *γ* = 2.5 – 3.0 of Fig. 3 leads to *β* = 2.0 – 3.0.

We note that as *β* → 2, our model becomes invalid as an exponential cutoff must be included. All figures are reproduced in the SI including such a cutoff. For *β* = 2.5 the numerical deviations of superspreading statistics are small when comparing the model with and without an exponential cutoff.

In Fig. 4(a) we show the classical superspreading plot: the fraction *f*_*x*_ of infected people infected by the *x* most infectious people. The concavity of these curves signify superspreading. The inset, adapted from Ref. [2], shows, for instance, that for the SARS epidemic, the 20 % most infectious infected more than 80 % of the total.

**FIG. 4.**
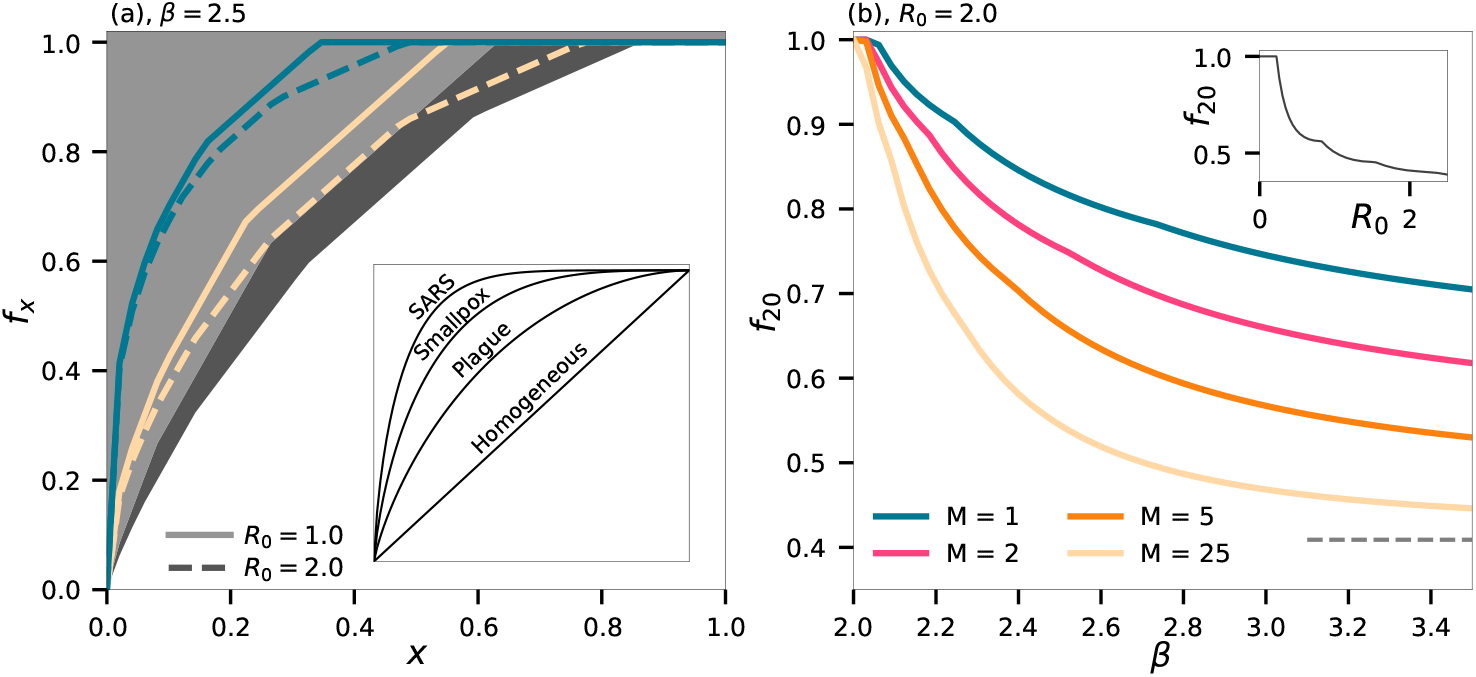
Spreading statistics. **(a)** The fraction *f*_*x*_ of people infected by the *x* most infectious people. Dark background indicates limit set by Poisson statistics alone. Colored indicate *M* as in (b), and full lines are for *R*_0_ = 1.0 and dashed for *R*_0_ = 2.0. Inset adapted from Ref. [2] showing estimated graphs for select historical epidemics. **(b)** The fraction of people *f*_20_ that the most infectious 20 % will infect for *R*_0_ = 2.5 as it varies with *β*. Inset shows *f*_20_ in the simple case of pure Poisson statistics as a function of *R*_0_. Dashed line in main plot indicates the Poisson value for *R*_0_ = 2.0.

In reality, the ‘ideal’ homogeneous distribution shown in the inset, in which the top *x* infectors infect exactly *x*, is unattainable simply due to the randomness of Poisson statistics. For finite *R*_0_, the grey (*R*_0_ = 1.0) and dark-grey (*R*_0_ = 2.0) background indicate the minimal statistics demanded by Poisson randomness alone. As we include spatial heterogeneity, the statistics become more extreme, especially for small *M* [blue curves, Fig. 4].

Fig. 4(b) fixes *x* = 20 % and the curves show *f*_20_, the fraction infected by the most infectious 20 % as a function of *β*. The inset shows the fraction demanded by Poisson statistics alone. For *R*_0_ = 2.0 and *β* = 2.5, the fraction infected by the most infectious 20 % depends strongly on *M*. For a short time of infectious, *M* = 1, we find *f*_20_ ≈ 85 %. Already at *M* = 2 this falls to *f*_20_ ≈ 75 %, and at *M* = 25 we find *f*_20_ ≈ 55 %. Thus if a disease has a short period of high infectiousnes it can show similar heterogeneous infection pattern as an epidemic dominated by biological superspreaders. As *β* is lowered, the statistics become more extreme, but so does the effect of an exponential cutoff. We show in the SI, that with a cutoff of *n* = 1,000, *f*_20_ can never be larger than 90 % (for *M* = 1).

### Mitigation

As an epidemic unfolds, mitigation measures can be employed to halt the spread of disease. In SIR models, effects of mitigation are captured by lowering the infectiousness. [*α* in our model]. In contrast, in network models [17] the number of connections between individuals can be lowered for a similar effect.

We capture the effects of mitigation by calculating how a measure affects the instantaneous value of *R*_0_. Mitigation measures such as enforcing the use of masks and good hygiene will directly affect *α*. As *R*_0_ is directly proportional to *α*, the effect of this measure is trivial. As we directly model people visiting different locations, we focus instead on the effect of closing locations. We emphasize that the effect on *R*_0_ of closing places is in fact independent on duration of infectiousness (*M*) in our model, and is thus effective even for a disease that does not show spreading heterogeneity. This is a direct consequence of allowing infectious individuals to infect many in each time step.

Instructionally, we begin by calculating the effect of closing locations of a given size. Fig. 5 shows that if small locations are closed, *R* is actually increased as some of the people that would visit a small location instead go to larger locations. However when locations of a reasonable size are closed, we see that *R* decreases. This effect subsequently diminishes for very large *n*, as there are only few large gatherings of a particular size.

**FIG. 5.**
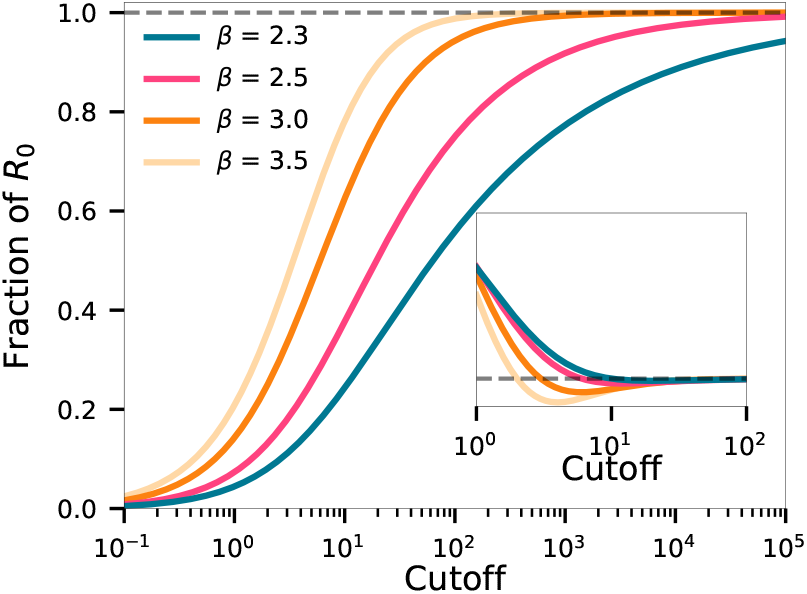
Effect of mitigation by banning gatherings. Curves shows the fractional change in *R*_0_ as a function of a cutoff size, larger than which gatherings are disallowed. Inset shows the effect of bans of a specific size, dashed line indicating the line of no change. Note that the cutoff can be fractional in that we consider the *average* number of interactions at a location.

A more natural approach is to close all places larger than a given cutoff size, i.e. a ban on large gatherings. We derive in the SI, the distribution resulting from replacing *P*_*N*_ (*n*) with 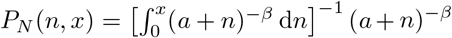, where *x* is the location cutoff size. Fig. 5 shows that a much stricter mitigation effort is required to bring down *R* for large *β* than for small. For small *β*, a large fraction of all infectious take place at large locations and thus one can get away with a smaller mitigation effort. It is thus crucial to gauge the value of *β* for a community when choosing the size of mitigation needed to bring *R*_0_ below one.

### Contact Tracing

A separate mitigation effort comes through contact tracing [18, 19]: finding and isolating people who have been in contact with an infectious individual. The duration of infectiousness *M* and the power law exponent *β* likewise have big consequences for the effectiveness of this approach.

We show in Fig. 6(a) the probability, given a person was infected, that he was infected at an event of size *z* ≥ 50. If this probability is high, the effect of contact tracing is eased, as the epidemic will be driven by large spreading events.

**FIG. 6.**
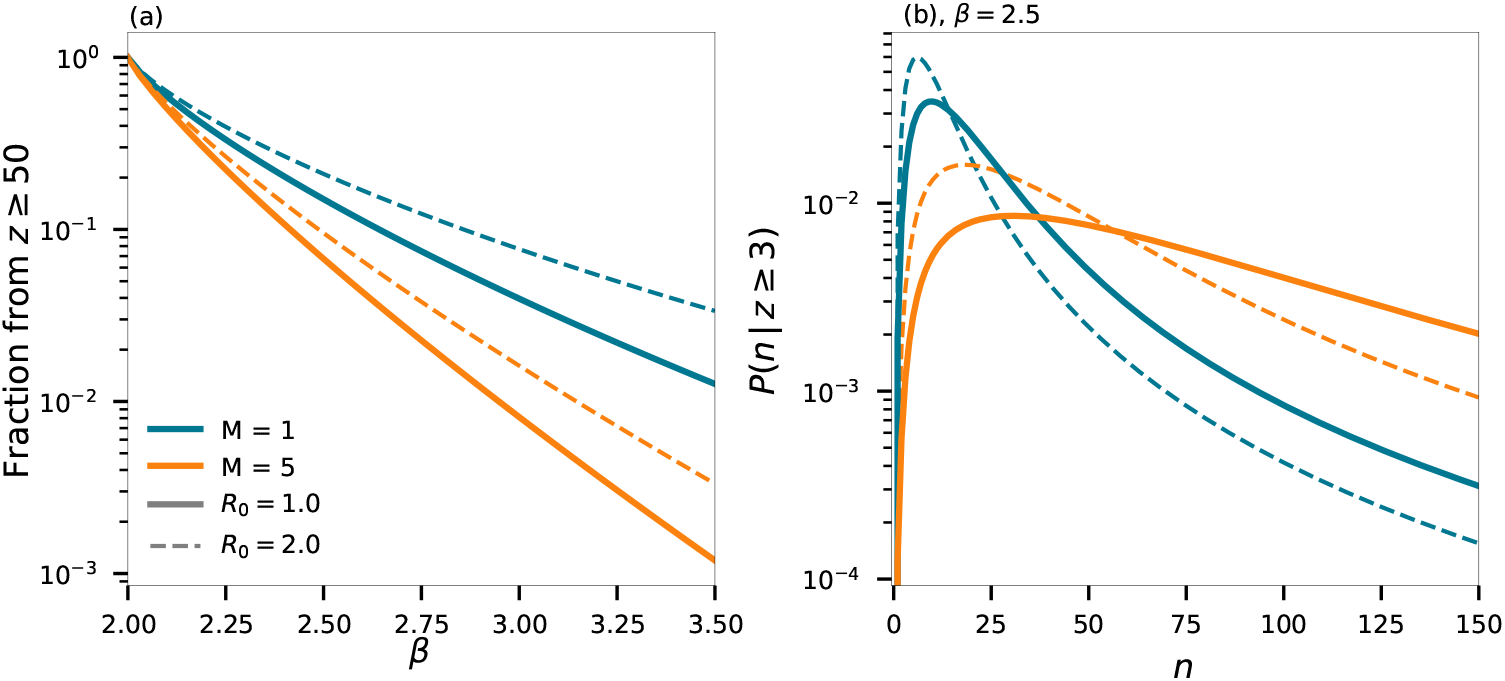
**(a)** The fraction of people infected in an event in which more than 50 people got infected as a function of *β*. **(b)** The probability density that 3 people, who were infected at the same infected, were infected at an event where they had *n* interactions on average. *M* and *R*_0_ vary as indicated by the legend of (a).

Our model further allows us to identify the likelihood of where events took place. For instance, consider the case where, say, *c* = 3 members of a family all got infected and we can assume that they got infected at the same event. Where is this most likely to have happened? This will depend on the socio-spatial parameters of the community they live in and on the duration of infectiousness of the disease. By the law of Bayes, we have

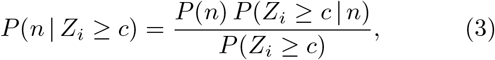

where the terms on the right side are given by Eq. (1), the partial sum of a Poisson distribution, and 1 minus the sum of Eq. (4), respectively. Note that this equation depends on *a* and *α* independently. Fig. 6(b) shows this for *c* = 3 and *β* = 2.5. We see that, for fixed *R*_0_, the larger the time of infectiousness, *M*, the more likely it is that this happened at a large location. In the same manner, the value of *R*_0_ affects the distribution as well as shown in the figure.

## DISCUSSION

We have presented a model that is simple enough to be studied analytically, yet reveals key insights into the statistics of epidemics spreading in heterogeneous spaces. The model is equally applicable to populations ranging from a full country to a small isolated neighbourhood, the difference being modelled by the parameter *β* and the exponential cutoff. In contrast, the infectiousness period *M* is set by the disease, and thus does not change by location.

Given knowledge of a disease and the space in which it is spreading, our model gives baseline statistics which can give quite extreme measures (e.g. 20 % infecting 80 %) even in the absence of biological superspreaders. We thus conclude that to judge directly from data if super-spreaders are important drivers in an epidemic, it should also be contrasted to a baseline model that include spatial heterogeneity and duration of infectiousness instead of a simple homogeneous population.

Our model does not directly take into account effects such as a variance in infectiousness attributed to factors such as ventilation or the type of event taking place at a location (concert or dinner?). However, this simply amounts to a re-interpretation of *P*_*N*_, as variations in the type of event would either increase or decrease the number of *effective* interactions.

In the SI we develop and solve analytically a version of the model that includes an exponential cutoff. Such a cutoff is necessary to get accurate statistics for *β* ≲ 2.5 (dependent on the exact cutoff). In particular, with an exponential cutoff we can further allow *β <* 2, which perhaps could be representative for very social demographics such as young people. The value of the exponential cut-off could be estimated based on spreading events: If one observes that it is rare to find a person who infects more than *k* other people, then one can employ an exponential cutoff in the order of *k/α*.

With an exponential cutoff, a bound can be put on statistics such as *f*_20_ even without knowing the value of *β*. Thus, by simply knowing the approximate value of the exponential cutoff as well as the duration of infectiousness, our model predicts extremeness statistics such as *f*_20_. If these are found to be more extreme in an ongoing epidemic it would be strong evidence for the existence of biological superspreaders no matter the value of *β*. For example, as detailed in the SI, if the exponential cut- off is *s* ∼ 1,000, we find for a disease with *R*_0_ = 2.0 that *f*_20_ ≳ 0.9 implies the existence of superspreaders for *M* = 1 and any value of *β*. For *M* = 5 this drops to *f*_20_ ≳ 0.7, while for *M* = 25 we have *f*_20_ ≳ 0.55. We note further that an alternative regularization to an exponential cutoff is a direct cap on the number of interactions. This is equivalent to a ban of gatherings, the details of which can be found in the SI.

While the above arguments show that knowledge of *β* is not always required, our model naturally gives more accurate results when *β* is known. We have presented example data for which *β* can be approximately extracted. The social mobility data is generated from Bluetooth signals between smartphones of ∼650 students. Naturally, these people will in general be in rooms with many other people, for whom we have no data. Thus the data will in general underestimate the number of people at locations. In particular, after study/work hours will be highly underestimated as people will go home or away from university campus, where there is little chance to encounter other students that participate in the study. Conversely, student life tend to quite regularly gather people in large crowds for instance at lectures. Thus depending on the range over which we extract the power law, different exponents are found. The data, in any case, demonstrates that power laws can approximate the size distribution of locations that people visit also on the scale of 10^2^ – 10^3^ people.

To pinpoint social heterogeneity our study calls for active sociological studies that goes beyond the contact time measurements of Ref. [20]. Most effectively this could be done by mobile tracking, in analogy to the limited study among student used in our work. With social structure input and estimates of *M* and *R*_0_ for a given ongoing disease we may fix *α*. The assumption of all people being equally infectious can then be tested from observed rate of household transmissions [21], taking into account the fraction of time spent by individuals in the household. Likewise, one of the parameters of the model could be fixed by comparing to e.g. the fraction of people that infect no one [Fig. 2].

In the context of the 2019 SARS-CoV-2 epidemic, there is strong evidence that the spread is by aerosols [22]. Further, viral load during the infection is quite peaked [23], which could indicate a short period of high infectiousness on the order of 1 – 2 days. Superspreaders have been suggested to be a large factor in driving the SARS-CoV-2 epidemic [6], which, in turn, has an effect on the choice of mitigation efforts. It is thus of paramount importance to fully understand the true distribution of superspreaders, taking into account that superspreading statistics can result from many sources, one being socio-spatial heterogeneity as discussed here.

Our model could further be expanded to explicitly include biological superspreaders. For instance, if one expects varying viral loads to be driving the variation in spreading, the constant infectivity *α* can be replaced with a dispersed probability distribution with mean value *α*. In this case, special care should be taken when *α* for some highly infectious individuals can exceed unity. Below this limit, however, our conclusions on mitigation strategies by banning large gatherings remain unaffected. If, on the other hand, variations in spreading are due to e.g. varying particle sizes of the airborne pathogens, the variation should instead be applied to *P*_*N*_ directly, as some individuals will be able to infect a higher fraction of people at a given location. For example, if only some people produce pathogenic aerosol these would have a much higher range than others [14]. In a location of *x* people, such a superspreader might reach the entirety of the *x* people, instead of just 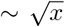. This would take the exponent *β* from, say, 2.5 to 1.75.

Our model further gives insights into applying mitigations. In the case of SARS-CoV-2, many countries chose to implement bans on large gatherings. These range from banning gatherings larger than 1,000 people e.g. in France, to banning social gatherings of more than 6 people in the U.K. Fig. 5 shows how the difference in effectiveness of such bans depend strongly on the value of *β*. In particular, the shape of the power law in Eq. (1) set the range over which changing the size of the ban has the highest effect. Naturally, the lower the *β* the better the effect of a ban of a certain size. However, one may also consider the effect of changing a ban from one size to another. For instance, for *β* = 2.5 changing from a ban of gathering larger than 100 to 10 reduces *R* by ∼ 50%, whereas for *β* = 3.0 the reduction is more than ∼ 68%. Further, as long as the ban size is significantly smaller than the exponential cutoff, the effect of such a cutoff is negligible.

The traditional approach to modelling a heterogeneous population is to consider network models. In our model, time is discretized by location visits, during which an infectious individual can, in principle, infect the entirety of an event. In contrast, in SIR and network models infectious individual typically interact with one person per time step. Our approach thus focuses on airborne pathogens, where transmission does not require direct touch, and has the effect that the number people that are together becomes central. This in turn leads to the prediction that mitigation strategies aimed at reducing *R*_0_ depends strongly on the socio-spatial distribution of gatherings, but is in fact independent of the duration of infectivity *M*. Thus we find that for airborne diseases, mitigation by reducing large gatherings will work well for diseases that do not exhibit heterogeneous spreading (*M* ≫ 1).

The distance of infection of such airborne transmissions can vary a lot between diseases and this should be reflected in the final choice of distribution exemplified by Eq. (1). In particular, for diseases that spread as aerosol, the distance of influence can become very big whereas a disease that is transmitted by larger droplets only allow limited secondary infections at even very large events. Likewise, in calculating the statistics of our model we have assumed a fully susceptible population. For diseases such as influenza, where cross-immunity plays a major role [24–26], the number of susceptible people at the social events should be re-scaled accordingly.

## METHODS

To derive the statistics of our model, we consider the early stages of an epidemic where only a fraction of the entire population has been infected. Here, we can neglect saturation and derive analytically the offspring distribution in terms of *α* and the social parameters *a* and *β* from Eq. 1. We find

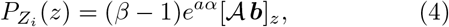

where 𝒜 is a matrix with entries

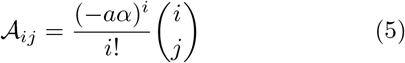

and ***b*** is a vector with entries

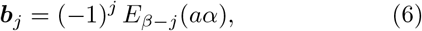

and *E*_*n*_(·) is the exponential integral function. We relegate the derivation to the SI, but note that Eq. (4) is exact even when truncating 𝒜 and ***b*** to be finite. We furthermore use 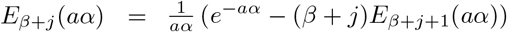, permitting evaluation by recursion as long as 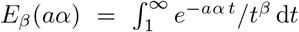 is known, which can be evaluated using Gaussian quadrature or any other standard method.

Finally, we can use a (zero-padded) discrete Fourier transform (DFT) to obtain the probability for any *M* :

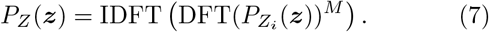

An important special case of our general formula is

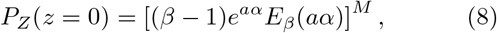

the probability that a person infects exactly zero after having visited *M* places. These people represent an important dual aspect of superspreading: When few are infecting the majority, only few infection events are left to the majority of people.

We note that *a* and *α* always appear in the combination *a* · *α*, thus one of them can be chosen freely if the other is adjusted to fix *R*_0_. In this way, our calculations are valid for any value of *a* as long as *α* is adjusted accordingly, and thus the exact behaviour of the power law at small values is not important for the statistics when considering fixed *R*_0_.

For the purpose of estimating the social parameter *β*, we utilized data collected from smartphones distributed to around 1,000 students at a Danish university [27]. Of these students we only used the subset of 642 students who had daily records of proximity to others. The cell phone tracking data consist of proximity measurements achieved from repeated Bluetooth scans by the smartphones every fifth minute. We divide the data into into blocks of one hour, and subsequently discretize the data in a consistent manner by capping the received signal strength indicator at a value that corresponds to ∼2 meters. This results in hourly networks of students that are within the vicinity of one another. We use the connected components of these graphs as indicators for the sizes of locations to generate Fig. 3(a). The relatively low number of total students who participate in the study combined with infrequent Bluetooth scans result in low exponential cutoffs for the number of direct connections within 2 meters. Nonetheless, as an approximation for the range of an airborne pathogens, we consider the average number of students that are directly connected within each cluster. Fig. 3(b) shows that this average approximately grows like the square root of the cluster size. The true number of potential infections in a location of a given size depends on the precise characteristics of the disease, in particular if it spreads as aerosol or not [14].

For estimating the amount of time people spent together at locations, we calculate the distribution of the duration of meetings between all pairs of students. Two students are considered to be in the same room, if they are in the same cluster. We only have Bluetooth signals every fifth minute, and a person can be beyond the reach of this signal for a brief time without having actually left a certain location. Thus we permit gaps in the tracks. Fig. 3 shows the result of allowing gaps of 5 minutes and 30 minutes. Naturally, the latter yields larger times spent together, but the difference is not massive. We further note that the data show peaks at around 2 hours and 4 hours, which we suspect to be the result of students going to a single lecture or two lectures in a row, respectively.

## Data Availability

Data available upon request.

## Acknowledgements

This project has received funding from the European Research Council (ERC) under the European Union’s Horizon 2020 Research and Innovation Programme, Grant Agreement No. 740704.

## References

[1] Richard A Stein. Super-spreaders in infectious diseases. International Journal of Infectious Diseases, 15(8):e510–e513, 2011.

[2] J. O. Lloyd-Smith, S. J. Schreiber, P. E. Kopp, and W. M. Getz. Superspreading and the effect of individual variation on disease emergence. Nature, 438(7066):355–359, 2005.

[3] Akira Endo, Sam Abbott, Adam J Kucharski, Sebastian Funk, et al. Estimating the overdispersion in covid-19 transmission using outbreak sizes outside china. Wellcome Open Research, 5(67):67, 2020.

[4] Danielle Miller, Michael A Martin, Noam Harel, Talia Kustin, Omer Tirosh, Moran Meir, Nadav Sorek, Shiraz Gefen-Halevi, Sharon Amit, Olesya Vorontsov, et al. Full genome viral sequences inform patterns of sars-cov-2 spread into and within israel. medRxiv, 2020.

[5] Dillon Adam, Peng Wu, Jessica Wong, Eric Lau, Tim Tsang, Simon Cauchemez, Gabriel Leung, and Benjamin Cowling. Clustering and superspreading potential of severe acute respiratory syndrome coronavirus 2 (sars-cov-2) infections in hong kong. 2020.

[6] Taylor Robert J. Sneppen, Kim and Lone Simonsen. Impact of Superspreaders on dissemination and mitigation of COVID-19. Medrxiv, page 2020.05.17.20104745, 2020.

[7] Thomas R Frieden and Christopher T Lee. Identifying and interrupting superspreading events—implications for control of severe acute respiratory syndrome coronavirus 2. 2020.

[8] Lea Hamner. High sars-cov-2 attack rate following exposure at a choir practice—skagit county, washington, march 2020. MMWR. Morbidity and Mortality Weekly Report, 69, 2020.

[9] Max SY Lau, Bryan Grenfell, Kristin Nelson, and Ben Lopman. Characterizing super-spreading events and age-specific infectivity of covid-19 transmission in georgia, usa. medRxiv, 2020.

[10] Shweta Bansal, Bryan T Grenfell, and Lauren Ancel Meyers. When individual behaviour matters: homogeneous and network models in epidemiology. Journal of The Royal Society Interface, 4(16):879–891, 2007.

[11] Bjarke Frost Nielsen, Kim Sneppen, Lone Simonsen, and Joachim Mathiesen. Heterogeneity is essential for contact tracing. medRxiv, 2020.

[12] Alexei V Tkachenko, Sergei Maslov, Ahmed Elbanna, George N Wong, Zachary J Weiner, and Nigel Goldenfeld. Persistent heterogeneity not short-term overdispersion determines herd immunity to covid-19. arXiv preprint 2008.08142, 2020.

[13] William Ogilvy Kermack, A. G. McKendrick, and Gilbert Thomas Walker. A contribution to the mathematical theory of epidemics. Proceedings of the Royal Society of London. Series A, Containing Papers of a Mathematical and Physical Character, 115(772):700–721, 1927.

[14] Martin Z. Bazant and John W. M. Bush. Beyond six feet: A guideline to limit indoor airborne transmission of covid-19. medRxiv, 2020.

[15] R. L. Axtell. Zipf distribution of U.S. firm sizes. Science, 293(5536):1818–1820, 2001.

[16] Xavier Gabaix. Power laws in economics: An introduction. Journal of Economic Perspectives, 30(1):185–206, 2016.

[17] Bjarke Frost Nielsen and Kim Sneppen. Covid-19 super-spreading suggests mitigation by social network modulation. medRxiv, 2020.

[18] Joel Hellewell, Sam Abbott, Amy Gimma, Nikos I Bosse, Christopher I Jarvis, Timothy W Russell, James D Munday, Adam J Kucharski, W John Edmunds, Fiona Sun, et al. Feasibility of controlling covid-19 outbreaks by isolation of cases and contacts. The Lancet Global Health, 2020.

[19] Andreas Eilersen and Kim Sneppen. Estimating cost-benefit of quarantine length for covid-19 mitigation. medRxiv, 2020.

[20] Joël Mossong, Niel Hens, Mark Jit, Philippe Beutels, Kari Auranen, Rafael Mikolajczyk, Marco Massari, Stefania Salmaso, Gianpaolo Scalia Tomba, Jacco Wallinga, et al. Social contacts and mixing patterns relevant to the spread of infectious diseases. PLoS Med, 5(3):e74, 2008.

[21] Wei Li, Bo Zhang, Jianhua Lu, Shihua Liu, Zhiqiang Chang, Cao Peng, Xinghua Liu, Peng Zhang, Yan Ling, Kaixiong Tao, and Jianying Chen. Characteristics of Household Transmission of COVID-19. Clinical Infectious Diseases, 04 2020. ciaa450.

[22] Kimberly A. Prather, Linsey C. Marr, Robert T. Schooley, Melissa A. McDiarmid, Mary E. Wilson, and Donald K. Milton. Airborne transmission of sars-cov-2. Science, 370(6514):303–304, 2020.

[23] Andrew W Byrne, David McEvoy, Aine Collins, Kevin Hunt, Miriam Casey, Ann Barber, Francis Butler, John Griffin, Elizabeth Lane, Conor McAloon, et al. Inferred duration of infectious period of sars-cov-2: rapid scoping review and analysis of available evidence for asymptomatic and symptomatic covid-19 cases. medRxiv, 2020.

[24] Derek J Smith, Alan S Lapedes, Jan C de Jong, Theo M Bestebroer, Guus F Rimmelzwaan, Albert DME Osterhaus, and Ron AM Fouchier. Mapping the antigenic and genetic evolution of influenza virus. science, 305(5682):371–376, 2004.

[25] Julia R Gog and Bryan T Grenfell. Dynamics and selection of many-strain pathogens. Proceedings of the National Academy of Sciences, 99(26):17209–17214, 2002.

[26] Florian Uekermann and Kim Sneppen. A cross-immunization model for the extinction of old influenza strains. Scientific reports, 6:25907, 2016.

[27] Arkadiusz Stopczynski, Vedran Sekara, Piotr Sapiezynski, Andrea Cuttone, Mette My Madsen, Jakob Eg Larsen, and Sune Lehmann. Measuring large-scale social networks with high resolution. PloS one, 9(4):e95978, 2014.

