## Supplemental Text for "Airborne Pathogens in a Heterogeneous World: Superspreading & Mitigation"

### Supplemental Information

Julius B. Kirkegaard, Joachim Mathiesen, and Kim Sneppen
*Niels Bohr Institute, University of Copenhagen, 2100 Copenhagen, Denmark*
(Dated: October 24, 2020)

#### Distribution

Marginalizing over

$$P_N(n) = (\beta - 1)\frac{a^{\beta-1}}{(a+n)^\beta} \tag{1}$$

$$P_{Z_i}(z) = \frac{(\alpha n)^z e^{-\alpha n}}{z!} \tag{2}$$

gives

$$P_Z(z) = \frac{(\beta-1)a^{\beta-1}}{z!}\int_0^\infty \frac{(\alpha n)^z e^{-\alpha n}}{(a+n)^\beta}\,\mathrm{d}n \equiv \frac{(\beta-1)a^{\beta-1}}{z!} f(z,\beta). \tag{3}$$

We only need the integral for integer values of $z$, so we may do the calculation recursively:

$$\begin{aligned} f(z+1,\beta) &= \int_0^\infty \frac{(\alpha n)^{z+1} e^{-\alpha n}}{(a+n)^\beta} \\ &= \alpha \int_0^\infty \frac{n(\alpha n)^z e^{-\alpha n}}{(a+n)^\beta} \\ &= \alpha\left[\int_0^\infty \frac{(a+n)(\alpha n)^z e^{-\alpha n}}{(a+n)^\beta} - a\int_0^\infty \frac{(\alpha n)^z e^{-\alpha n}}{(a+n)^\beta}\right] \\ &= \alpha\left[\int_0^\infty \frac{(\alpha n)^z e^{-\alpha n}}{(a+n)^{\beta-1}} - a\int_0^\infty \frac{(\alpha n)^z e^{-\alpha n}}{(a+n)^\beta}\right] \\ &= \alpha\left[f(z,\beta-1) - a f(z,\beta)\right]. \end{aligned} \tag{4}$$

With this recursion, all values $0 \le z \le n$ can be calculated in $\mathcal{O}(n^2)$ evaluations if we know $f(0,\beta)$. Luckily this integral is much simpler:

$$f(0,\beta) = e^{a\alpha}\,\alpha^{\beta-1}\,\Gamma(1-\beta, a\alpha) = e^{a\alpha}\,a^{1-\beta}\,E_\beta(a\alpha), \tag{5}$$

where $\Gamma(\cdot,\cdot)$ is the incomplete gamma function and $E_n(\cdot)$ is the exponential integral function. Finally we solve the recursion to obtain the result of the main text.

#### Exponential Distribution

When the method of disease transmission is by physical touch, the exponential distribution becomes more relevant. Taking

$$P_N(n) = s^{-1}e^{-n/s} \tag{6}$$

we find

$$P_{Z_i}(z) = (\alpha s)^z\,(1+\alpha s)^{-(z+1)}. \tag{7}$$

In this case we can directly solve for the distribution for any $M$. Calculating the probability generating function, we find

$$G_{Z_i}(\zeta) = \frac{1}{1+\alpha s(1-\zeta)}. \tag{8}$$

We raise this to the $M$-th power and take $z$ derivatives to find

$$P_Z(z) = \binom{M+z-1}{z} \frac{(\alpha s)^z}{(1+\alpha s)^{M+z}}. \tag{9}$$

This has average $\alpha M s$, which for fixed $R_0$ implies $\alpha = R_0/(sM)$. Thus the distribution is independent of $s$ for fixed $R_0$:

$$P_Z(z) = \binom{M+z-1}{z} \frac{(R_0/M)^z}{(1+R_0/M)^{M+z}}. \tag{10}$$

This is a negative binomial distribution, $\mathrm{NB}(M, (1+R_0/M)^{-1})$. It corresponds to the superspreading distribution of Ref. [1] if one identifies their dispersion parameter $k$ with $M$. The correspondence arises because the sum of $M$ exponentially distributed random numbers is gamma distributed and the sum of Poisson distributed random numbers is itself Poisson distributed.

Since the distribution is independent of $s$, so be its output statistics. In particular, the above distribution gives $f_{20} \approx 0.60$ for $M = 1$ and $R_0 = 2.0$, whereas we have $f_{20} \approx 0.65$ for $R_0 = 1.0$. For $M = 5$ the fractions fall to $f_{20} \approx 0.46$ ($R_0 = 2.0$) and $f_{20} \approx 0.54$ ($R_0 = 1.0$). Since these are valid for all $s$, we conclude that extreme statistics such as 20 % infecting 80 % can never be achieved from exponential distributions.

**Mitigation**

For mitigation we redistribute people below a maximum $n = x$. So we have

$$P_N(n) = \frac{1}{\int_0^x \frac{1}{(a+n)^\beta}\,\mathrm{d}n} \frac{1}{(a+n)^\beta} \equiv \frac{1}{N(a,\beta,x)} \frac{1}{(a+n)^\beta} \tag{11}$$

where the normalization is a simple, but ugly integral. We now need

$$P_Z(z) = \frac{1}{z!N(a,\beta,x)} \int_0^x \frac{(\alpha n)^z e^{-\alpha n}}{(a+n)^\beta}\,\mathrm{d}n \equiv \frac{1}{z!N(a,\beta,x)} f_x(z,\beta). \tag{12}$$

By the exact same argument as above, we have

$$f_x(z+1,\beta) = \alpha\left[f_x(z,\beta-1) - a f_x(z,\beta)\right]. \tag{13}$$

Again we need the value for $z = 0$, which in this case is

$$f_x(0,\beta) = e^{a\alpha}\,\alpha^{\beta-1}\left(\Gamma(1-\beta, a\alpha) - \Gamma(1-\beta,(a+x)\alpha)\right). \tag{14}$$

.

**Exponential Cutoff**

We use a power law for the distribution of sizes of location. This creates two problems: (1) the model becomes invalid as $\beta \to 2$ and (2) higher order moments diverge. To fix these issues an exponential cutoff must be included. In this case we consider

$$P_N(n) = \frac{a^{\beta-1}}{E_\beta(a/s)} \frac{e^{-\frac{a+n}{s}}}{(a+n)^\beta}, \tag{15}$$

where $s$ denotes the (soft) cutoff value. Now we have

$$P_Z(z) = \frac{a^{\beta-1}}{z!E_\beta(a/s)} \int_0^\infty \frac{(\alpha n)^z e^{-\alpha n} e^{-\frac{a+n}{s}}}{(a+n)^\beta}\,\mathrm{d}n \equiv \frac{a^{\beta-1}}{z!E_\beta(a/s)} f_s(z,\beta), \tag{16}$$

and again

$$f_s(z+1,\beta) = \alpha\left[f_s(z,\beta-1) - a f_s(z,\beta)\right], \tag{17}$$

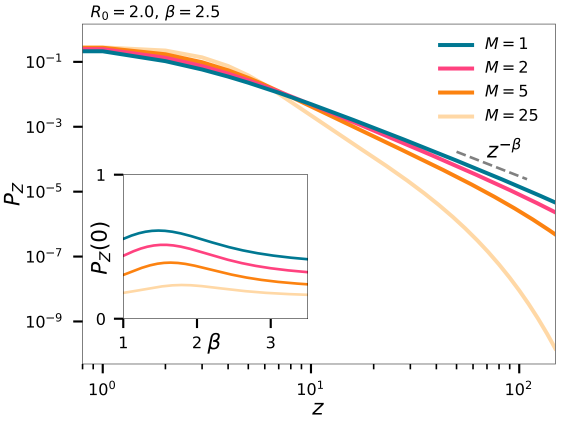


FIG. 1. Same as main text, but with exponential cutoff $s = 1{,}000$.

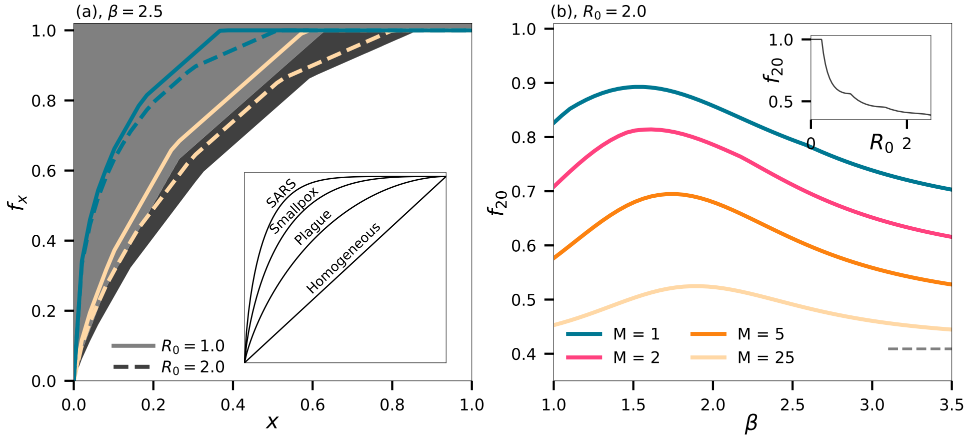


FIG. 2. Same as main text, but with exponential cutoff $s = 1{,}000$.

where now

$$f_s(0, \beta) = e^{a\alpha}\, a^{1-\beta}\, E_\beta(a\alpha + as^{-1}). \tag{18}$$

Solving this recursion leads to

$$P_Z(z) = \frac{(-a\alpha)^z e^{a\alpha}}{z!\, E_\beta(a/s)} \sum_{j=0}^{z} (-1)^j \binom{z}{j} E_{\beta-j}(a\alpha + as^{-1}). \tag{19}$$

For $\beta \geq 1$, we have $1/E_\beta(0) = (\beta - 1)$, confirming that this result reduces to that of the main text for $s \to \infty$. Note that for $s$ finite, we no longer need to require $\beta > 2.0$. Fig. 3 shows the plot $f_{20}$ with an exponential cutoff. We note that the values of $\beta = 2.5$ are virtually unchanged. Interestingly, the curves now yield a maximum. Thus, with a cutoff, the superspreading statistics emerging from spatial heterogeneity can be bounded even if $\beta$ is unknown.

#### Mitigation & Exponential Cutoff

We now have

$$P_N(n) = \frac{1}{\int_0^x \frac{e^{-n/s}}{(a+n)^\beta}\, \mathrm{d}n} \frac{e^{-n/s}}{(a+n)^\beta} \equiv \frac{1}{N(a, \beta, x)} \frac{e^{-n/s}}{(a+n)^\beta}. \tag{20}$$

As before this leads us to

$$P_Z(z) = \frac{1}{z! N(a, \beta, x)} \int_0^x \frac{(\alpha n)^z e^{-n/s} e^{-\alpha n}}{(a+n)^\beta}\, \mathrm{d}n \equiv \frac{1}{z! N(a, \beta, x)} f_x(z, \beta). \tag{21}$$

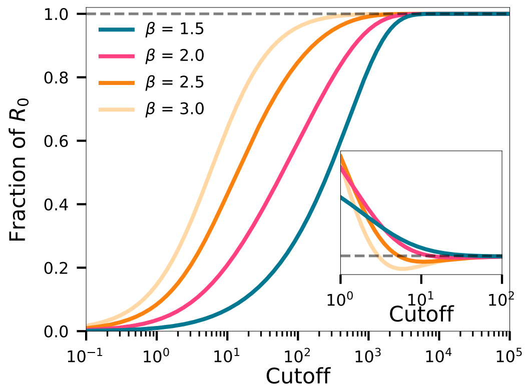


FIG. 3. Same as main text, but with exponential cutoff $s = 1{,}000$.

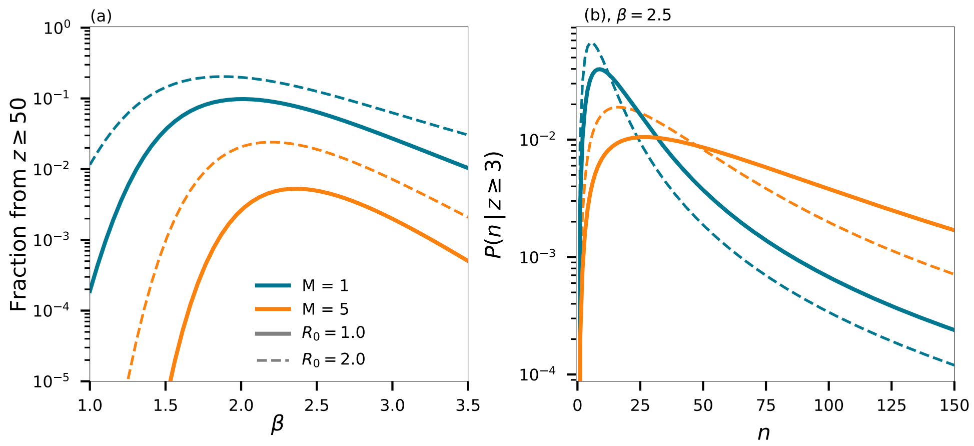


FIG. 4. Same as main text, but with exponential cutoff $s = 1{,}000$.

By the exact same argument as above, we have

$$f_x(z+1,\beta) = \alpha\left[f_x(z,\beta-1) - af_x(z,\beta)\right]. \tag{22}$$

Again we need the value for $z = 0$, which in this case is

$$f_x(0,\beta) = \frac{s\,(\alpha+s^{-1})^{\beta}e^{a\alpha+as^{-1}}}{1+\alpha s}\left[\Gamma(1-\beta,a\alpha+as^{-1}) - \Gamma(1-\beta,(a+x)(1+s\alpha)s^{-1})\right]. \tag{23}$$

.

[1] J. O. Lloyd-Smith, S. J. Schreiber, P. E. Kopp and W. M. Getz. Superspreading and the effect of individual variation on disease emergence. *Nature*, 438, 355–35, 2005.
